# ICU admission and mortality in adult patients with influenza A/H1N1-related pneumonia in Vietnam since the 2009 H1N1 pandemic: a 10-year cohort study

**DOI:** 10.64898/2026.04.18.26351156

**Authors:** Quang Minh Ho, Bich Thuy Duong, Le Nhu Tung Nguyen, Tri Nugraha Susilawati, Bui Ngoc Minh Tam, Thanh Truc Thai, David J. Muscatello, Anthony Sunjaya, Siyu Chen, Thanh Nguyen Nguyen, Minh Thu Nguyen, Kim Anh Nguyen, Minh Cuong Duong

## Abstract

The A(H1N1)pdm09 virus remains a major global health threat. This study examined the burden of ICU admission, mortality, and associated predictors among patients with A(H1N1)pdm09 pneumonia in a leading center for infectious diseases in Vietnam. Information on demographic, clinical, and laboratory characteristics, and outcomes was retrieved from medical records of adults admitted with influenza A(H1N1)pdm09 during 2009–2019. Among 729 cases, 21.7% (158/729) developed pneumonia. Among 158 pneumonia cases, 36.7% (58/158) developed moderate-to-severe acute respiratory distress syndrome (ARDS), and 15.2% (24/158) received invasive ventilation. ICU admission and mortality rates were 48.7% (77/158, 95%CI 41.1–56.5%) and 8.2% (13/158, 95%CI 4.9–13.6%), respectively. Predictors of ICU admission included being >60 years old (adjusted OR [AOR] 13.864, 95%CI 2.185–87.956, P=0.005), comorbidities (AOR 6.527, 95%CI 1.710–24.915, P=0.006), AST (AOR 1.013, 95%CI 1.001–1.025, P=0.029), and moderate-to-severe ARDS (AOR 14.027, 95%CI 4.220–46.627, P<0.001). Predictors of mortality were invasive ventilation (AOR 55.355, 95%CI 1.486–2062.375, P=0.030) and double-dose oseltamivir or combination therapy (AOR 32.625, 95%CI 1.594–667.661, P=0.024). In conclusion, mortality is not rare in A(H1N1)pdm09 infection. Monitoring of older patients and those with comorbidities, liver enzyme elevation, or moderate-to-severe ARDS is essential for the timely detection of complications requiring intensive care.

## Introduction

The A(H1N1)pdm09 virus emerged in 2009 and triggered a global influenza pandemic associated with an estimated 123,000–203,000 deaths (1, 2). The pandemic was declared over in August 2010, after which A(H1N1)pdm09 transitioned into an endemic seasonal strain co-circulating with influenza A(H3N2) and B viruses (3, 4). While influenza seasonality is well defined in temperate regions, circulation patterns in tropical areas are often irregular, which may compromise the optimal timing and impact of vaccination programs (5). Despite the availability of effective vaccines, A(H1N1)pdm09 remains a significant contributor to worldwide influenza-related morbidity and mortality (6). A study conducted in China found that during 2010 and 2015, the death associated with influenza was 9.9 per 100,000 people, in which influenza A(H3N2) virus was the leading cause with a rate of 5.18 per 100,000 followed by A/(H1N1)pdm09 with a rate of 2.9 per 100,000 (7).

While most influenza infections resolve without treatment, some patients, especially high-risk groups, may develop serious complications that can lead to death(3). High-risk groups include children younger than five years, the elderly, pregnant women, and individuals with compromised immune systems or chronic medical conditions (8–10).However, the literature reports inconsistent risk factors for severe outcomes, which may partly explain the observed between-country differences in A(H1N1)pdm09 mortality burden (11, 12). A study involving 696 A/(H1N1)pdm09 patients in Spain has concurred that immune deficiency and chronic cardiovascular diseases are risk factors for mortality (10). Interestingly, this Spanish study has found that older age is associated with lower ICU admission but a risk factor for death, suggesting age may play a differential role in predicting ICU admission versus mortality (10). Regarding pregnancy, given that this is a well-accepted risk factor for severity and mortality of A/(H1N1)pdm09 pneumonia, the World Health Organization (WHO) recommends that pregnant women are prioritized for influenza vaccination (13). In contrast to this established understanding, a Mexican study[MH1.1][MH1.2][DBT(1.3] involving 2,944 A/(H1N1)pdm09 patients has found a reduced fatality rate among pregnant women, showing variations in the association of pregnancy and disease severity from A/H1N1 infections in different countries (11).

The dissimilarities in identifying risk factors for A/(H1N1)pdm09 pneumonia-related mortality worldwide may reflect differences in study design, timing, population age structures, ethnic composition, geographical settings, and vaccination coverage (14). Frequent viral mutations that influence disease severity may further contribute to variability in findings (15). Therefore, ongoing, robust, country-specific studies are needed to improve understanding of the risk factors of A/(H1N1)pdm09 infection-related ICU admission and mortality. However, long-term influenza surveillance remains limited in tropical regions of the Asia-Pacific due to gaps in information technology and available resources, with surveillance systems largely focusing on basic epidemiological data but lacking clinical and outcome information (16, 17). Most studies in these areas are short-term (typically 1–2 years) (18), which is insufficient for effective monitoring, as influenza circulation in tropical settings tends to be sporadic and unpredictable. Establishing a comprehensive, nationwide surveillance system requires substantial financial investment, trained personnel, and infrastructure capable of collecting and integrating individual-level data. Moreover, because many countries in the region face competing public health priorities (19), maintaining long-term influenza surveillance systems is often challenging and may not be feasible or sustainable (18, 20). In Vietnam, influenza viruses co-circulated all year round and were an important cause of influenza-like illness between 2006 and 2010 (21). The first H1N1 vaccine became available locally in 2010, followed by other vaccines targeting both seasonal influenza strains and A(H1N1)pdm09. However, these vaccines are accessible primarily on a self-paid basis, even for high-risk groups. To strengthen influenza prevention and control in Vietnam and similar settings, continual assessment of disease severity is needed (21). This study examined the mortality burden and other treatment outcomes, as well as identified risk factors for ICU admission and mortality, among adult patients suffering from A/(H1N1)pdm09 pneumonia in Vietnam.

## Materials and Methods

### Study design and context

A 10-year retrospective cohort study was conducted at the Hospital for Tropical Diseases (HTD) in Ho Chi Minh City from 1st January 2009 to 31st December 2019. The study was approved by the Ethics Committee of HTD (approval number 43/HĐĐĐ). HTD is the 550-bed referral hospital for infectious diseases in southern Vietnam, receiving patients from across this region, including the Mekong Delta, with approximately 3,000 patients accessing healthcare services daily (22). HTD’s medical records of hospitalized patients are standardized in accordance with the Vietnam Ministry of Health’s guidelines (23).

All hospitalized adult patients diagnosed with A/(H1N1)pdm09 pneumonia during the study period were enrolled in the study. The inclusion criteria included persons aged 17 years and older (referred to here as ‘adults’), having a positive reverse transcription polymerase chain reaction (RT-PCR) result with A/(H1N1)pdm09 from nasal or oropharyngeal swab, and developing pneumonia confirmed by chest x-rays. There were no exclusion criteria.

### A(H1N1)pdm09 laboratory confirmation, definitions of A(H1N1)pdm09 pneumonia, and indications for ICU admission

Nasal and oropharyngeal swabs were collected by treating doctors and nurses as per the HTD’s guidelines. The RT-PCR was performed on the LightCycler 480 II System (Roche Molecular Diagnostics, Pleasanton, CA) (24, 25) using the QIAGEN OneStep RT-PCR kit (25, 26). The 20 µl of reaction volume was prepared and included 5 µl of extracted RNA template, 0.5 µl of enzyme mix, 10 µl of 2× reaction mix, 0.25 µmol/L influenza A forward primer, 0.2 µmol/L influenza A reverse primer, 0.25 µmol/L influenza A probe, 0.25 µmol/L influenza B forward primer, 0.2 µmol/L influenza B reverse primer, and 0.25 µmol/L influenza B original probe. This process had been validated elsewhere (27). All laboratory tests were performed at the standardized Laboratory Department (ISO 15189) of the HTD.

Patients with A/(H1N1)pdm09 pneumonia were defined as those who had lung injuries confirmed by chest x-rays demonstrating parenchymal and/or interstitial injuries or alveolar injuries. Indications for ICU admission were in accordance with the national guidelines for diagnosis and treatment of influenza (28) and international recommendations (9, 29)including development of respiratory distress or septic shock. Respiratory distress is defined as difficulties in breathing or shortness of breath, PaO2/FIO2 ≤200 mmHg or SpO2/FIO2 ≤235 (if SpO2 ≤97%) (30). Septic shock is identified with a clinical construct of sepsis with persisting hypotension requiring vasopressors to maintain MAP ≥65 mmHg and having a serum lactate level >2 mmol/L (18 mg/dL) despite adequate volume resuscitation (31).

### Data collection

A data collection form was used to record participants’ information from HTD’s paper-based medical records. Data included demographics (age, gender, comorbidity, and pregnancy status of females), clinical signs and symptoms, chest x-rays features, laboratory test results, treatments, complications, and outcomes. Clinical features included days from onset to hospital admission, fever, fatigue, cough, dyspnea, sore throat, chest pain, and moderate-to-severe acute respiratory distress syndrome (ARDS). Based on the Berlin definition, ARDS was defined as an acute disorder that starts within seven days of the inciting event and is characterized by bilateral lung infiltrates and severe progressive hypoxemia in the absence of any evidence of cardiogenic pulmonary edema (30). ARDS was categorized as mild, moderate, and severe (30). Chest x-ray features included unilateral or bilateral lung infiltrates and alveolar injuries. Laboratory characteristics included complete blood count, serum creatinine, alanine transaminase (ALT), aspartate transaminase (AST), total bilirubin, glycemia, serum albumin, C-reactive protein (CRP), procalcitonin, and arterial lactate. Treatments included antiviral therapy (standard-dose [75mg/12h] or double-dose [150mg/12h] oseltamivir or combination therapy [oseltamivir plus zanamivir or ribavirin]), other medications (corticosteroids and vasopressor drugs), and respiratory support (nasal cannula, mask, high-flow nasal cannula (HFNC), continuous positive airway pressure (CPAP), invasive or prone ventilation). Complications included hospital-acquired infections defined as any infection including ventilator-acquired pneumonia (VAP), urinary tract infection and bloodstream infection developing after two days of admission. Outcomes included mortality, hospital discharge, inter-hospital transfer together with reasons, ICU and hospital length of stay.

### Statistical analysis

Data were managed and analyzed using SPSS software version 26. Categorical variables were presented as a count and percentage and compared using the Chi-squared test, while continuous variables were presented as mean ± standard deviation (SD) and compared using Student’s t-test. For comparison purposes, 95% confidence intervals (CIs) of the point incidence of A/(H1N1)pdm09-related pneumonia, ICU admission, mortality, discharge, and inter-hospital transfer were calculated. Logistic regression models were developed to test predictors of ICU admission and mortality related to severe A/(H1N1)pdm09 pneumonia. Given the potential for confounding among covariates, the purposeful selection process was used to identify covariates for the regression models. Following the methodology described elsewhere (32, 33), a more generous P-value cutoff of <0.25 in the univariate analysis was used. This approach was chosen because traditional cutoffs (such as 0.05) often fail to identify variables that, while not independently significant, act as important confounders or become significant when adjusted for other covariates. Furthermore, variables deemed clinically important based on the authors’ expertise were retained in the model regardless of their statistical significance, ensuring that the final model is both statistically robust and biologically plausible. Variables included in the model for predictors of ICU admission were gender; age >60 years old; presence of any comorbidity; day from onset to admission; white blood cell (WBC), neutrophil, lymphocyte, and platelet cell counts; serum creatinine; AST; ALT; and moderate-to-severe ARDS. Variables included in the model for predictors of mortality were gender, age >60 years old, presence of any comorbidity, day from onset to admission, WBC and lymphocyte counts, moderate-to-severe ARDS, invasive ventilation, double-dose oseltamivir or combination, corticosteroids, vasopressors, hospital-acquired infection, and ICU admission. Alpha was set at 5% level.

## Results

### Baseline characteristics of study participants

A total of 729 A(H1N1)pdm09 infected patients received treatment at the HTD during the study period (Figure 1). Most (73%, 532/729) patients were hospitalized in 2009 (Figure 2). Of these 729 patients, A(H1N1)pdm09-related pneumonia was diagnosed in 21.7% (158/729, 95% CI 18.8-24.8%).

**Figure 1.**
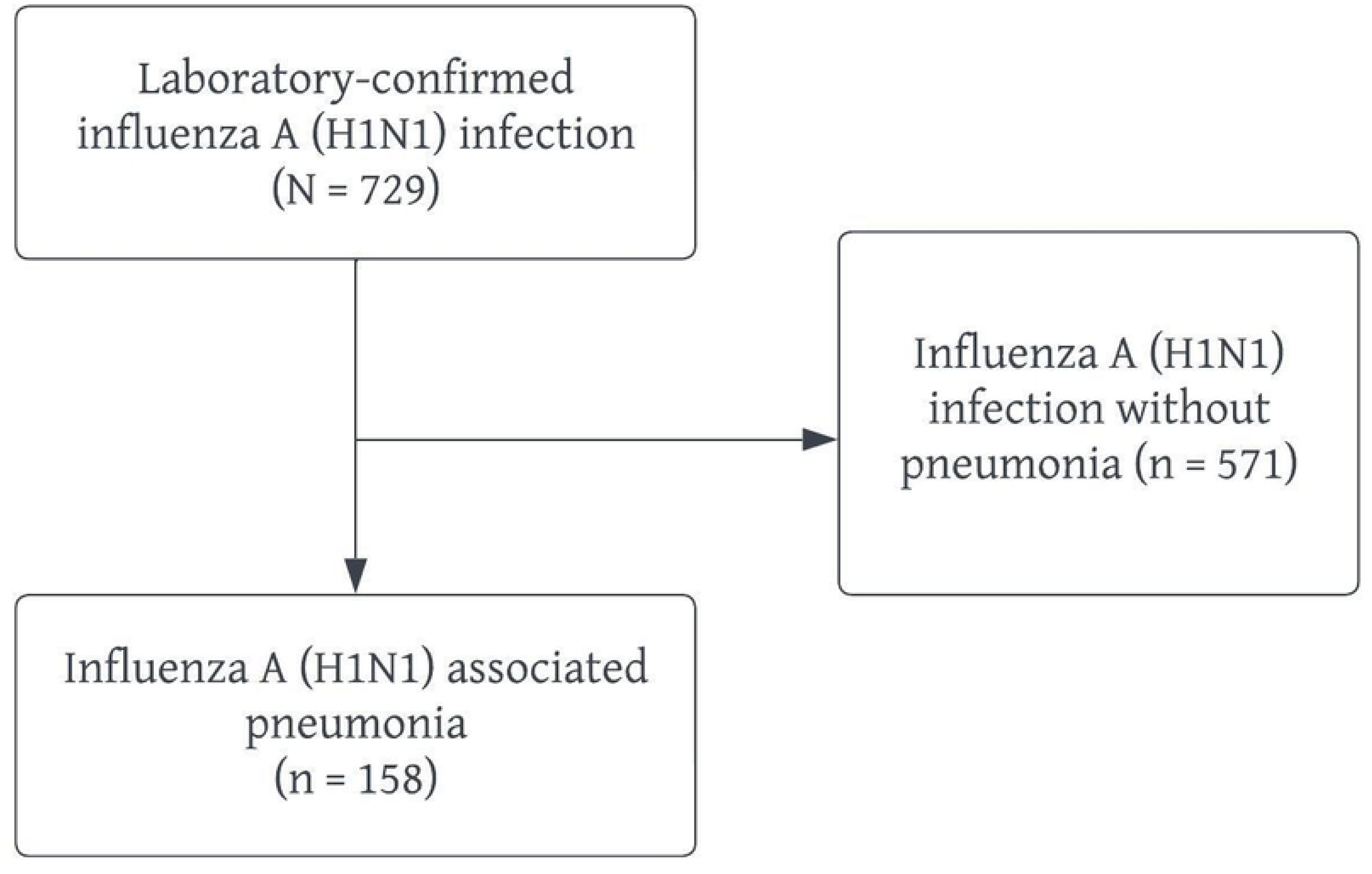
Flowchart of study participants.

**Figure 2.**
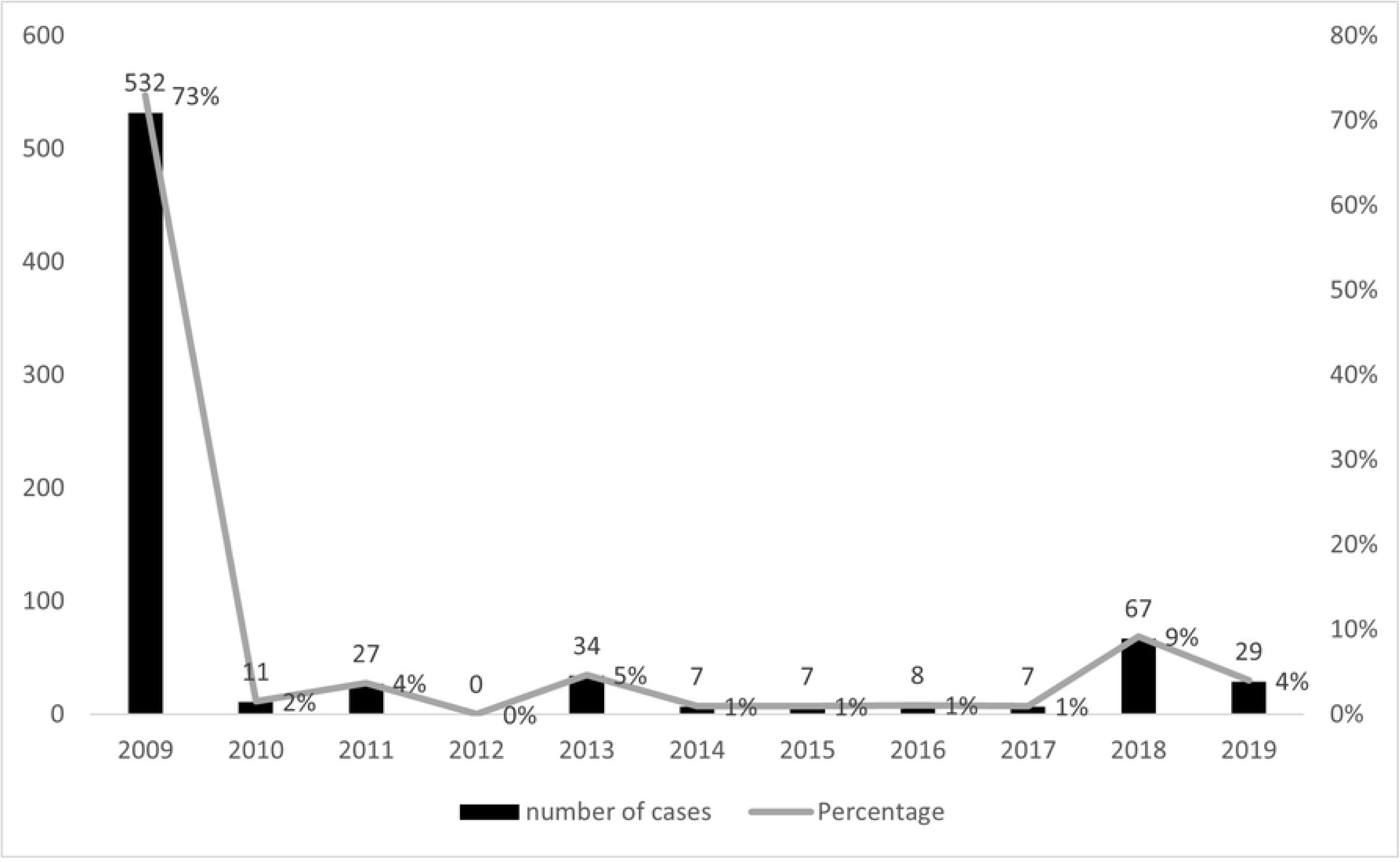
Frequency of laboratory-confirmed A(H1N1)pdm09 infected cases by year between 2009 and 2019.

The mean age of 158 participants with A(H1N1)pdm09-related pneumonia was 41.8 ± 16 years old, with 24 cases (15.2%) older than 60 years old. Among these 158 participants, 50.6% (80/158) were male, 16.5% (26/158) were pregnant, and 30.4% (48/158) had at least one comorbidity, including cardiovascular disease, diabetes, chronic liver disease, chronic lung disease (chronic obstructive pulmonary disease and asthma), and chronic kidney disease. The mean period from symptom onset to hospital admission was 4 ± 3 days. The most common presenting symptoms included fever (92.4%, 146/158), fatigue (61.4%, 97/158), and productive cough (51.9%, 82/158). Patients with moderate-to-severe ARDS accounted for 36.7% (58/158). Alveolar injuries (96.8%, 153/158) were the most common finding on chest x-rays (Table 1). Among the 26 pregnant patients included, 57.7% required ICU admission and three (11.5%) died. Four women went into labor while being treated for influenza, and one case resulted in stillbirth (data not shown).

**Table 1.**
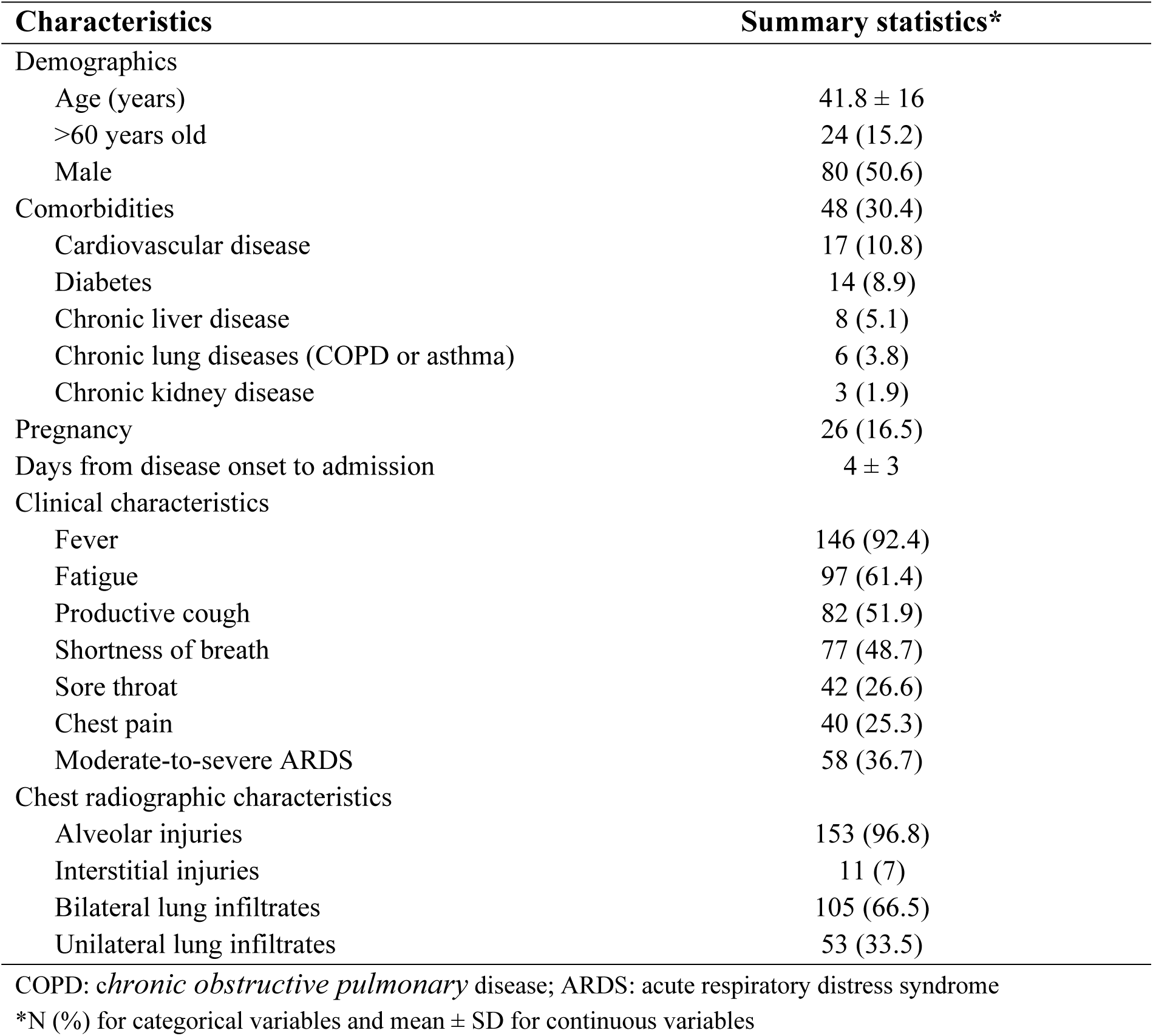
Demographic, clinical, and chest radiographic characteristics of 158 patients with laboratory-confirmed A(H1N1)pdm09-related pneumonia.

### Treatment and outcomes of study participants

Most patients (88%, 139/158) were treated with standard-dose oseltamivir. ICU admission accounted for 48.7% (77/158, 95%CI 41.1–56.5%). Just less than two-thirds (61.4%, 97/158) of patients received respiratory support, and invasive ventilation accounted for 15.2% (24/158). Hospital-acquired infections were documented in 6.3% (10/158) of patients. Most patients were discharged (86.1%, 136/158, 95%CI 79.8–90.6%), and 7.6% required inter-hospital transfer (12/158, 95%CI 4.4–12.8%). Reasons for inter-hospital transfer included treatment of tuberculosis and other comorbidities, and delivery services for pregnancy (data not shown). The mortality rate was 8.2% (13/158, 95%CI 4.9–13.6%). The mean hospital and ICU lengths of stay were 12.1 ± 9.1 and 6.8 ± 6.5 days, respectively (Table 2).

**Table 2.**
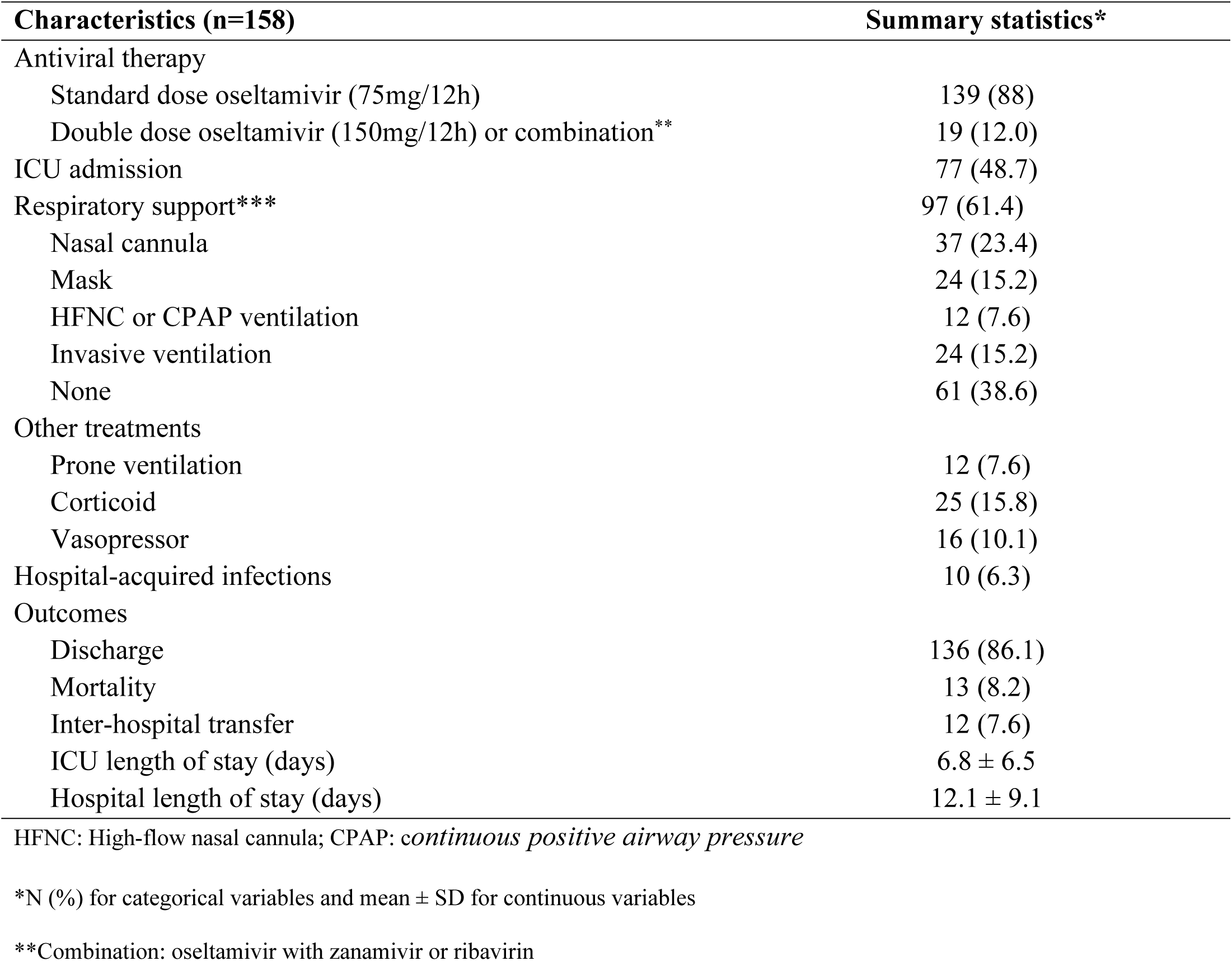
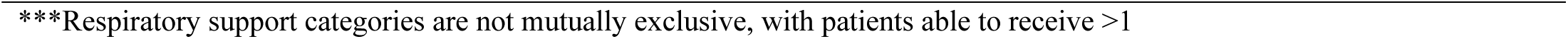
Antiviral and supportive therapies, complications, and treatment outcomes of 158 patients with A(H1N1)pdm09-related pneumonia.

| Characteristics (n=158) | Summary statistics* |
| --- | --- |
| Antiviral therapy |  |
| Standard dose oseltamivir (75mg/12h) | 139 (88) |
| Double dose oseltamivir (150mg/12h) or combination** | 19 (12.0) |
| ICU admission | 77 (48.7) |
| Respiratory support*** | 97 (61.4) |
| Nasal cannula | 37 (23.4) |
| Mask | 24 (15.2) |
| HFNC or CPAP ventilation | 12 (7.6) |
| Invasive ventilation | 24 (15.2) |
| None | 61 (38.6) |
| Other treatments |  |
| Prone ventilation | 12 (7.6) |
| Corticoid | 25 (15.8) |
| Vasopressor | 16 (10.1) |
| Hospital-acquired infections | 10 (6.3) |
| Outcomes |  |
| Discharge | 136 (86.1) |
| Mortality | 13 (8.2) |
| Inter-hospital transfer | 12 (7.6) |
| ICU length of stay (days) | $6.8 \pm 6.5$ |
| Hospital length of stay (days) | $12.1 \pm 9.1$ |

HFNC: High-flow nasal cannula; CPAP: *continuous positive airway pressure*
\*N (%) for categorical variables and mean ± SD for continuous variables
\*\*Combination: oseltamivir with zanamivir or ribavirin
\*\*\*Respiratory support categories are not mutually exclusive, with patients able to receive >1

### Unadjusted risk factors for ICU admission and mortality among 158 patients with laboratory-confirmed A(H1N1)pdm09-related pneumonia

There was a statistically significant association between ICU admission and comorbidities (OR = 1.812, 95%CI 1.348–2.435, P<0.001), moderate-to-severe ARDS (OR = 20.736, 95%CI 8.367–51.394, P<0.001), and laboratory parameters including serum creatinine (OR = 1.024, 95%CI 1.010–1.038, P<0.001), glycemia (OR = 1.007, 95%CI 1.001–1.014, P=0.026), AST (OR = 1.007, 95%CI 1.002–1.012, P=0.005), and serum albumin (OR = 0.775, 95%CI 0.654–0.918, P=0.003) (Table 3).

**Table 3.**
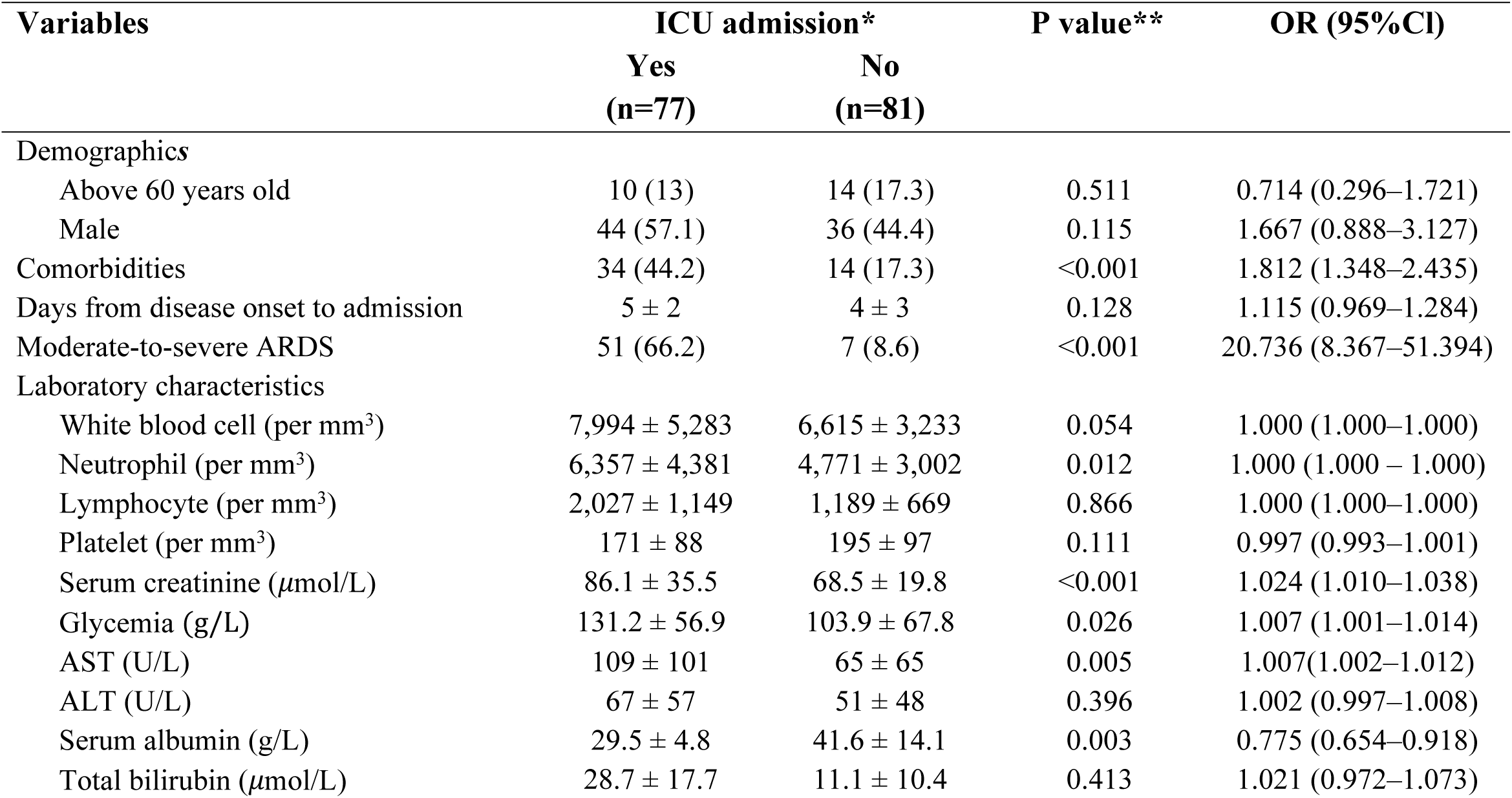
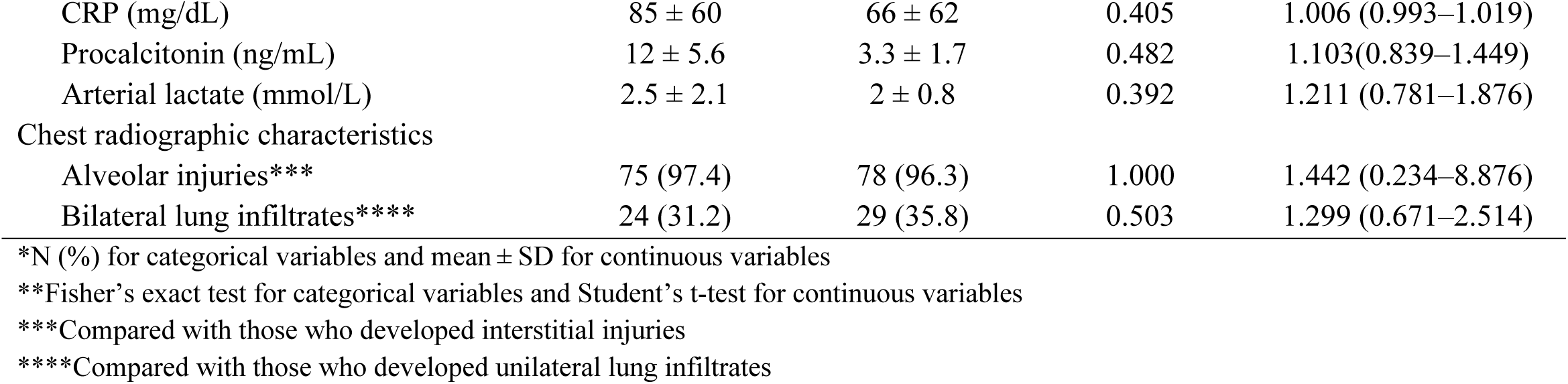
Unadjusted risk factors tested for ICU admission among 158 patients with laboratory-confirmed A(H1N1)pdm09-related pneumonia.

There was a statistically significant association between mortality and moderate-to-severe ARDS (P<0.001), laboratory parameters including AST (OR = 1.007, 95%CI 1.002–1.012, P=0.004) and treatment including the use of double-dose oseltamivir or combination (OR = 3.316, 95%CI 1.135–9.693, P=0.039) as well as invasive ventilation (OR = 47.143, 95%CI 9.376–237.039, P<0.001). There was also a statistically significant association between mortality and the use of corticosteroids (OR = 4.500, 95%CI 1.301–15.563, P=0.024) and vasopressors (OR = 8.766, 95%CI 2.385–32.225, P=0.003), hospital-acquired infections (OR = 7.400, 95%CI 2.681–20.422, P=0.003), and ICU admission (OR = 13.333, 95%CI 1.678–105.971, P=0.002) (Table 4).

**Table 4.**
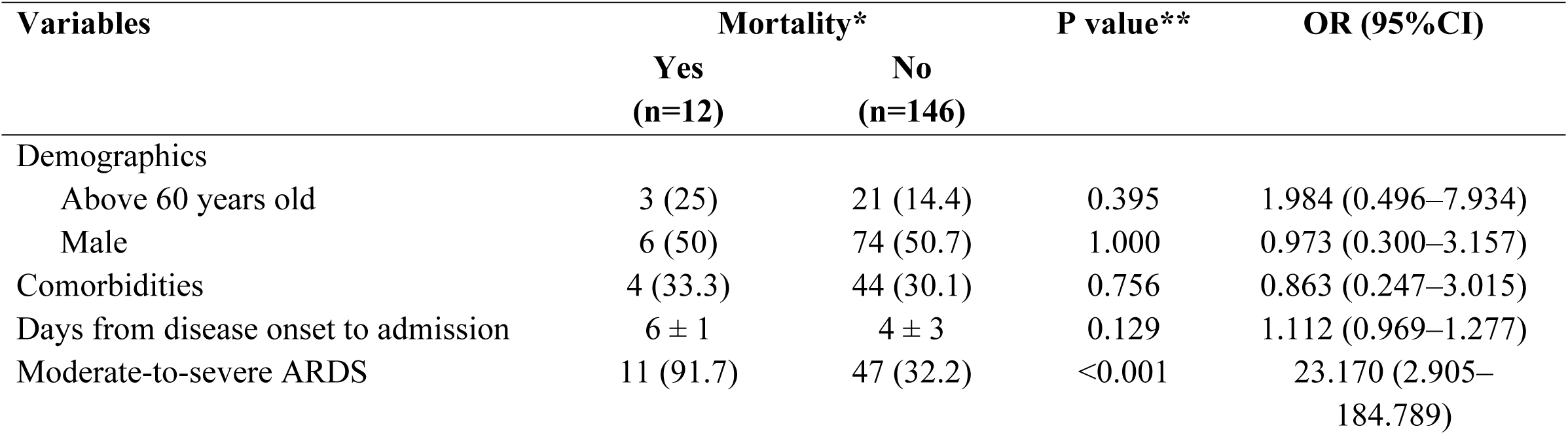
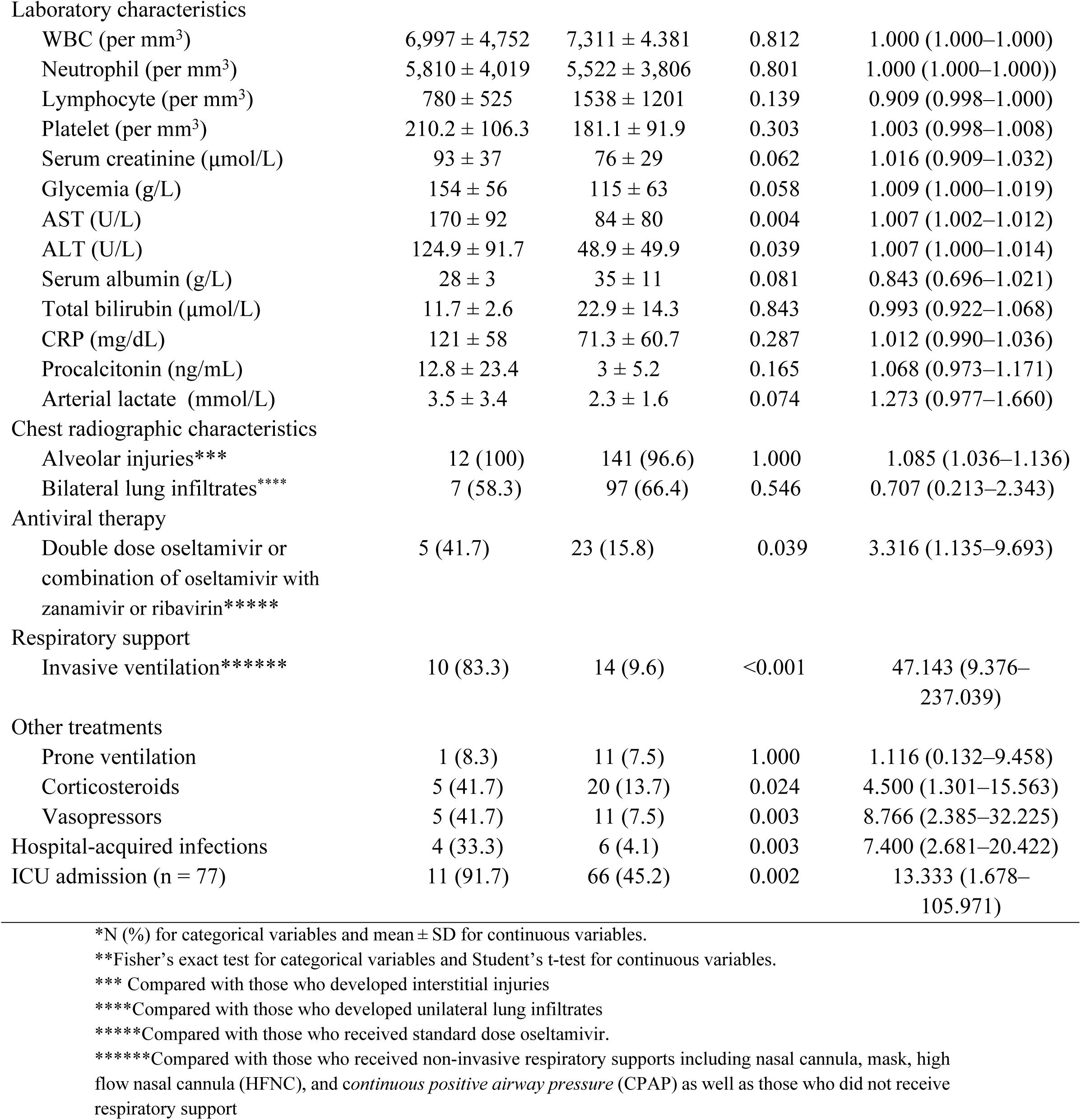
Unadjusted risk factors tested for mortality among 158 patients with laboratory-confirmed A(H1N1)pdm09-related pneumonia.

### Models for the prediction of ICU admission and mortality among 158 patients with laboratory-confirmed A(H1N1)pdm09-related pneumonia

Predictors of ICU admission included being above 60 years old (adjusted OR [AOR] 13.864, 95%CI 2.185–87.956, P=0.005), presence of any recorded comorbidity (AOR 6.527, 95%CI 1.710–24.915, P=0.006), AST (AOR 1.013, 95%CI 1.001–1.025, P=0.029), and moderate-to-severe ARDS (AOR 14.027, 95%CI 4.220–46.627, P < 0.001) (Table 5).

**Table 5.**
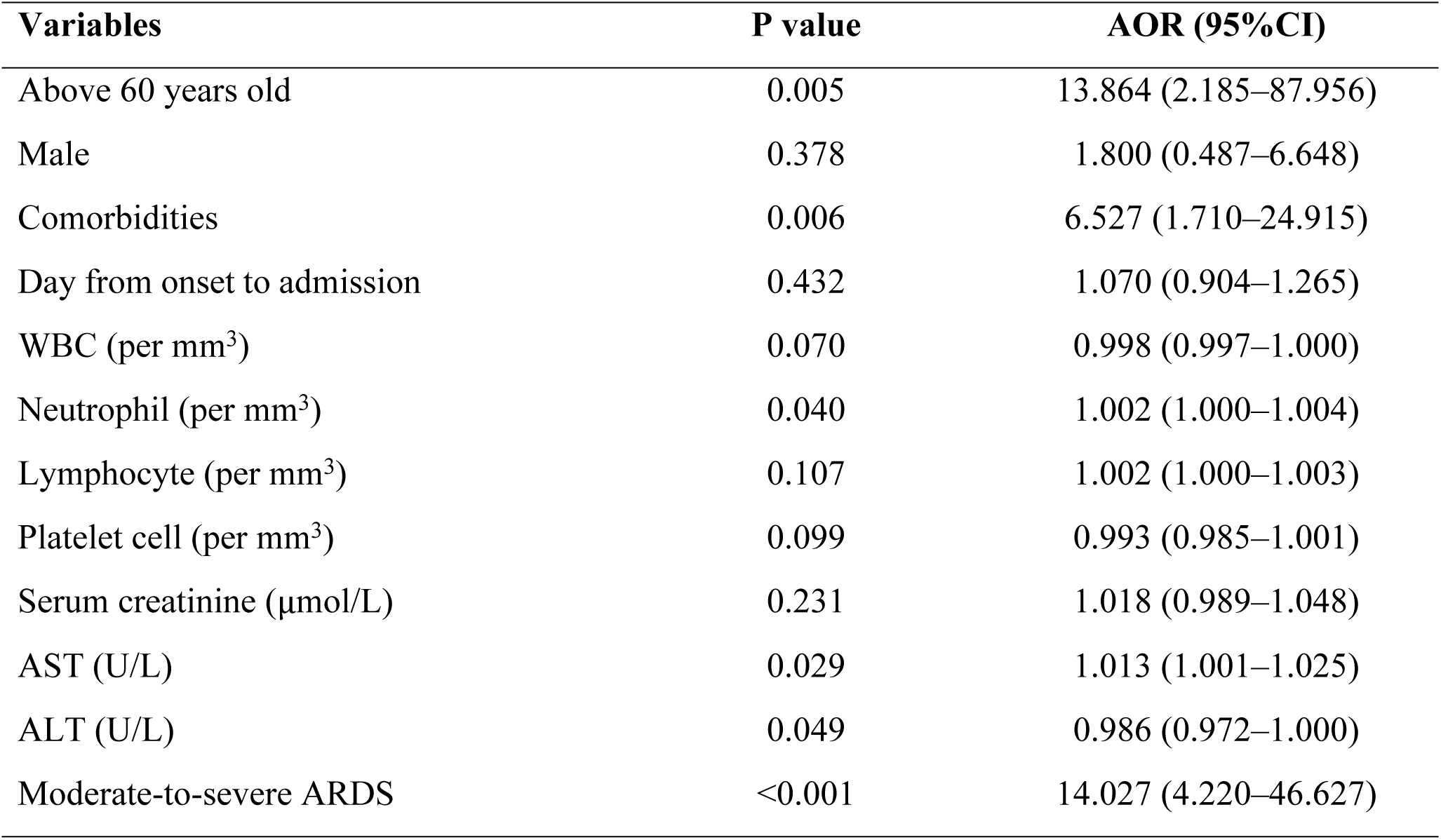
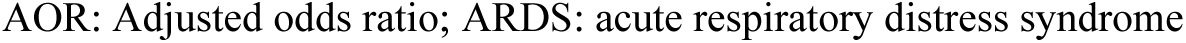
Logistic regression analysis of predictors of ICU admission among 158 patients with laboratory-confirmed A(H1N1)pdm09pneumonia.

| Variables | P value | AOR (95%CI) |
| --- | --- | --- |
| Above 60 years old | 0.005 | 13.864 (2.185–87.956) |
| Male | 0.378 | 1.800 (0.487–6.648) |
| Comorbidities | 0.006 | 6.527 (1.710–24.915) |
| Day from onset to admission | 0.432 | 1.070 (0.904–1.265) |
| WBC (per mm<sup>3</sup>) | 0.070 | 0.998 (0.997–1.000) |
| Neutrophil (per mm<sup>3</sup>) | 0.040 | 1.002 (1.000–1.004) |
| Lymphocyte (per mm<sup>3</sup>) | 0.107 | 1.002 (1.000–1.003) |
| Platelet cell (per mm<sup>3</sup>) | 0.099 | 0.993 (0.985–1.001) |
| Serum creatinine (μmol/L) | 0.231 | 1.018 (0.989–1.048) |
| AST (U/L) | 0.029 | 1.013 (1.001–1.025) |
| ALT (U/L) | 0.049 | 0.986 (0.972–1.000) |
| Moderate-to-severe ARDS | <0.001 | 14.027 (4.220–46.627) |

AOR: Adjusted odds ratio; ARDS: acute respiratory distress syndrome

Predictors of mortality included invasive ventilation (AOR 55.355, 95%CI 1.486–2062.375, P=0.030) and using double-dose oseltamivir or combination therapy (AOR 32.625, 95%CI 1.594–667.661, P=0.024) (Table 6).

**Table 6.** Logistic regression analysis of predictors of mortality among 158 patients with laboratory-confirmed A(H1N1)pdm09 pneumonia.

| Variables | P value | AOR (95%CI) |
| --- | --- | --- |
| Above 60 years old | 0.333 | 6.254 (0.153–255.87) |
| Male | 0.852 | 1.294 (0.087–19.356) |
| Comorbidities | 0.960 | 1.078 (0.059–19.571) |
| Days from onset to admission | 0.147 | 1.224 (0.931–1.610) |
| WBC (per mm<sup>3</sup>) | 0.484 | 1.000 (0.999–1.000) |
| Lymphocyte (per mm<sup>3</sup>) | 0.550 | 0.999 (0.996–1.002) |
| Serum creatinine (μmol/L) | 0.391 | 1.014 (0.982–1.046) |
| AST (U/L) | 0.530 | 1.006 (0.987–1.026) |
| ALT (U/L) | 0.995 | 1.000 (0.980–1.021) |
| Moderate-to-severe ARDS | 0.516 | 3.885 (0.065–233.589) |
| Invasive ventilation | 0.030 | 55.355 (1.486–2062.375) |
| Double dose oseltamivir (150mg/12h) or combination* | 0.024 | 32.625 (1.594–667.661) |
| Corticosteroids | 0.111 | 0.119 (0.009–1.633) |
| Vasopressors | 0.829 | 1.340 (0.093–19.231) |
| Hospital acquired infection | 0.506 | 2.849 (0.131–62.171) |
| ICU admission | 0.882 | 1.415 (0.02–136.79) |

AOR: Adjusted odds ratio; ARDS: acute respiratory distress syndrome
\*Combination: Oseltamivir with Zanamivir or Ribavirin

## Discussion

This study included all eligible patients treated at the HTD over a period of 10 years since the H1N1 pandemic. As the leading infectious disease center in southern Vietnam, HTD enabled this to be the largest and longest cohort study of its kind conducted in the country and likely in the wider tropical regions of the Asia-Pacific. Despite available treatments and vaccines, the study confirms a notable mortality in patients with A/(H1N1)pdm09-related pneumonia. To reduce mortality, older patients and those with comorbidities, elevated liver enzymes, or moderate-to-severe ARDS require close monitoring to detect complications early. Notably, rather than relying on double-dose oseltamivir or combination therapy, clinicians should prioritize early diagnosis, timely antiviral treatment, and appropriate antibiotics for confirmed bacterial co-infections.

There were 729 inpatients with A/(H1N1)pdm09 infection at the HTD during the study period. Of these patients, more than two-thirds were documented in 2009 when A/(H1N1)pdm09 emerged (3). Since then, A/(H1N1)pdm09 has continued to circulate as a seasonal flu and been included in the seasonal influenza vaccines (4). Given that influenza circulates year-round in Vietnam, both Southern and Northern hemisphere influenza vaccines together with a locally manufactured seasonal influenza vaccine are licensed (34). Information on the uptake of seasonal influenza vaccine in Vietnam is limited, and among the available literature on this topic, most studies were conducted on specific groups including health students and healthcare workers (34). Nevertheless, a local study conducted on 750 people in the general community found that 30% of participants received a seasonal flu vaccine either in the current or preceding flu season, and 64% expressed demand for this vaccine (35). Considering this, the decrease in the number of our A/(H1N1)pdm09 infected inpatients over time is probably attributable to the flu vaccination and herd immunity (36). Currently, flu vaccines are not part of the country’s broader immunization efforts (35). Considering the significant role of flu vaccination in preventing the disease, it is beneficial to add the vaccine to the Vietnam’s expanded immunization program, initially for priority populations with high risks of increased ICU admission and mortality.

Approximately 30% of our participants had at least one comorbidity, predominantly cardiovascular diseases and diabetes, and 16.5% were pregnant women. These underlying health conditions may exacerbate the severity of A/(H1N1)pdm09 infection (8). Although flu-like symptoms such as fever, cough, and sore throat are not specific for early diagnosis (37), most of our participants developed these symptoms. This finding was supported by previous studies indicating that flu-like symptoms can account for up to 100% of influenza patients (38). Alveolar injuries were the most common characteristics of lung injuries caused by A/(H1N1)pdm09 among our participants, predominantly representing bilateral lung infiltrates. A Japanese study similarly found that 71% (17/24) of their influenza patients developed bilateral lung injuries (39). However, several studies have indicated that the chest radiological findings of A/(H1N1)pdm09 infected patients are diverse and atypical (40), which may resemble those of other respiratory diseases, such as COVID-19 (40). Therefore, we believe that in clinical practice, a combination of thorough history-taking, clinical examination, and laboratory results is pivotal in making an accurate diagnosis and determining appropriate treatment.

Regarding treatment, all study participants were hospitalized early and treated with oseltamivir. A meta-analysis has shown that early treatment with oseltamivir can reduce the risk of lower respiratory tract complications and antibiotic usage in adult patients (41). Our participants demanded respiratory support at different levels, including non-invasive and invasive methods. Indeed, treatment for respiratory failure in patients with A/(H1N1)pdm09 pneumonia is similar to that of pneumonia caused by other agents and is based on patients’ severity of hypoxemia and underlying conditions (28). Despite effective treatment and vaccines, our study confirms that the mortality due to A/(H1N1)pdm09-related pneumonia is not rare (8.2%; 95% CI: 4.9–13.6%). It is comparable to that of a study conducted in India (8543/114667, 7.5%, 95% CI 7.3–7.6%) during a similar time period (42). In contrast, our mortality is lower than that of a study conducted in Singapore (41/172, 23.8%, 95%CI 18.1–30.7%) between June 2009 and August 2010 (43). Although our study period is longer than the Singaporean study, most of our patients were similarly recorded in 2009. The reasons for the differences in mortality rates are not clear and may warrant further studies. Nevertheless, the mortality rate due to A/(H1N1)pdm09 infection varies between countries, although it may be lower in developed countries, due to history of vaccination, differences in climate patterns, population density, and population age structure, as well as countries’ economic status and development (44). In Vietnam, our mortality rate is similar to that of a study conducted in the North of the country during 2015-2017 (25/259, 9.3%, 95%CI 6.0–13.7%) (45). In light of our findings, mortality due to A/(H1N1)pdm09-related pneumonia is not rare, despite effective treatment and vaccine

We found that those aged above 60 years old and with comorbidities are associated with an increased risk of ICU admission. Previous research similarly showed that elderly patients and those with comorbidities are associated with higher ICU admission rates, likely due to the increased risk of complications from A/(H1N1)pdm09 infection (3, 46). Other studies also found an elevation of both AST and ALT in several A/(H1N1)pdm09 infected patients, probably due to liver affection in the pathogenesis of influenza (47). Therefore, disease severity could adversely affect the liver function that subsequently causes elevated liver enzymes among patients admitted to ICU. We similarly observed an association between AST levels and ICU admission among our participants. Additionally, we also found that moderate-to-severe ARDS increases the risk of ICU admission. Considering our findings, patients with A/(H1N1)pdm09 infection who have at least one of following factors including advanced age, comorbidities, liver enzyme elevation, or moderate-to-severe ARDS should be monitored closely and continuously for timely detection of complications and the need for ICU admission. Indeed, at HTD, these characteristics have been trialed as screening criteria since the time the study was conducted. Our observations suggest that they are effective in identifying high-risk patients. Nevertheless, future studies are needed to validate the effectiveness and generalizability of these screening strategies. Regarding pregnancy, it was not identified as a predictor of disease severity or hospitalization in our study, likely due to the small number of pregnant participants. However, several complications were documented in this group. These findings highlight the substantial risks that influenza infection poses during pregnancy and underscore the importance of influenza vaccination in this high-risk group.

We found that mortality is strongly associated with invasive ventilation, which is consistent with findings from other studies (38, 48). Indeed, not all patients with influenza-related pneumonia undergo invasive ventilation, but those with more severe complications, including ARDS, are often given this type of ventilation support within 3–10 days following initial symptom onset (49). However, mechanical ventilation can worsen existing lung damage or even cause new lung injuries (50). Thus, studies are ongoing to identify the feasible and protective ventilation protocol for patients with influenza-related pneumonia requiring mechanical ventilation (50). Additionally, a study in China (51) has found that invasive ventilation is a risk factor for secondary bacterial pneumonia which can subsequently increase the risk of mortality among influenza patients. Although our findings did not demonstrate a significant relationship between hospital-acquired infections and mortality, we believe it is crucial to implement infection control and prevention strategies in critically ill patients with A/(H1N1)pdm09 infection on mechanical ventilation. We found that 12% of our study participants received oseltamivir at a double dose or in combination with other antiviral agents. However, we also found that double-dose oseltamivir or combination therapy was strongly associated with mortality. A recent systematic review and meta-analysis and another cohort study have shown that double-dose oseltamivir therapy does not help decrease mortality in patients with A/(H1N1)pdm09 infection (52, 53). However, this review found that in the subgroups of“the multicenter group” and“the large sample size group (n ≥ 200)”, the use of double-dose oseltamivir may be associated with an increased mortality with a low degree of heterogeneity (52). This is because most of the studies included in this review were observational and non-randomized, in which clinicians tended to prescribe higher doses of oseltamivir for severely ill patients who already had a higher risk of mortality (52). Regarding combination therapy, a recent study found that the combination of antiviral agents with different action mechanisms shortens the duration of shedding of viable virus in hospitalized patients with severe influenza but does not induce improved clinical outcomes (54). However, definitive evidence of the increased mortality risk associated with combination therapy is lacking. Given that our study is an observational study, we believe that there was also a tendency in prescribing combination therapy for patients with severe influenza, who were at a higher risk of mortality at our study site. Antiviral resistance was not assessed in this study. However, the potential risk of resistance, together with our findings on combination therapy, supports the conclusion of previous research (54). Based on our findings, the use of alternative antiviral regimens should not be recommended. Instead, priority should be given to improving early diagnosis, ensuring timely antiviral treatment, and using appropriate antimicrobials in patients with confirmed secondary bacterial infection, rather than initiating double-dose oseltamivir or combination antiviral therapy (54). We also did not find any significant association between age over 60 and mortality, which has been documented in other studies (55). Indeed, a large systematic review and meta-analysis found that the risk estimate for severe outcomes associated with influenza was heterogeneous in different countries classified by income levels (56). Therefore, we believe that more robust studies are needed to quantify the association between age and A/(H1N1)pdm09-associated mortality in Vietnam and comparable countries. These studies are also needed to fully understand the impact of double-dose oseltamivir and combination therapy on A/(H1N1)pdm09-associated mortality.

Our study has some notable limitations. Firstly, this study was based on an analysis of medical records and thus, data such as patients’ BMI (57) that may confound the study findings have not been documented. Secondly, our sample size was relatively small, which may have limited our ability to comprehensively investigate predictors of ICU admission and mortality in patients infected with A/(H1N1)pdm09. However, the dataset includes all eligible patients treated at the Hospital for Tropical Diseases between 2009 and 2019, providing a decade of real-world, post-pandemic data. Despite its modest size, the long-term dataset offers valuable longitudinal insights into disease patterns and clinical outcomes. Furthermore, the mortality rate over a 10-year period can be influenced by various factors such as improvement in hospital infrastructure and quality of medical service which can introduce bias into our analysis. Another limitation is that antiviral resistance was not assessed, which restricts our ability to fully evaluate the impact of double-dose oseltamivir and combination therapy on mortality risk. Future studies should include resistance testing to clarify this relationship. Evaluating the effectiveness of antibiotic therapy in patients with secondary infections represents an important avenue for future research but could not be conducted in our study due to the small number of confirmed cases. Nevertheless, the irregular circulation of A(H1N1)pdm09 in tropical regions and its ongoing antigenic changes underscore the need for continued surveillance. Our study provides updated evidence on the clinical features, laboratory findings, mortality patterns, and predictors of severe A(H1N1)pdm09 pneumonia in a tropical setting.

## Conclusions

Despite effective treatment and vaccines, our study confirms that the mortality due to A/(H1N1)pdm09-related pneumonia is not rare. In clinical practice, a thorough history-taking combined with clinical examination and laboratory results is essential for making an accurate diagnosis. Close and continuous monitoring of older patients and those with comorbidities, liver enzyme elevation, or moderate-to-severe ARDS is needed for a timely detection of complications that require intensive care and thus, reduce mortality. Neither double-dose oseltamivir nor combination therapy is recommended for the management of severe disease. Instead, clinical focus should be directed toward improving early diagnosis, ensuring the timely initiation of antiviral therapy, and maintaining appropriate antimicrobial stewardship when secondary bacterial infections are confirmed. Further robust, large-scale studies are warranted to more comprehensively profile mortality risk factors associated with A/(H1N1)pdm09 pneumonia in Vietnam and similar Asian populations.

## Data Availability

All relevant data are within the manuscript and its Supporting Information files.

## Acknowledgements

Not applicable.

## Supporting information

**S1 Fig**. Flowchart of study participants.

**S2 Fig**. Frequency of laboratory-confirmed A(H1N1)pdm09 cases by year between 2009 and 2019.

**S1 Table**. Demographic, clinical, and chest radiographic characteristics of 158 patients with laboratory-confirmed A(H1N1)pdm09-related pneumonia.

**S2 Table**. Antiviral and supportive therapies, complications, and treatment outcomes of 158 patients with A/(H1N1)pdm09-related pneumonia

**S3 Table**. Unadjusted risk factors tested for ICU admission among 158 patients with laboratory-confirmed A/(H1N1)pdm09-related pneumonia

**S4 Table**. Unadjusted risk factors tested for mortality among 158 patients with laboratory-confirmed A/(H1N1)pdm09-related pneumonia

**S5 Table**. Logistic regression analysis of predictors of ICU admission among 158 patients with laboratory-confirmed A/(H1N1)pdm09pneumonia

**S6 Table**. Logistic regression analysis of predictors of mortality among 158 patients with laboratory-confirmed A/(H1N1)pdm09 pneumonia

**List of abbreviations**
ALT: alanine transaminase
ARDS: acute respiratory distress syndrome
AST: aspartate transaminase
Cis: confidence intervals
CPAP: continuous positive airway pressure
CRP: C-reactive protein
HFNC: high-flow nasal cannula
HTD: Hospital for Tropical Diseases
ICU: intensive care unit
RT-PCR: reverse transcription polymerase chain reaction
SD: standard deviation
VAP: ventilator-acquired pneumonia
WBC: white blood cell
WHO: World Health Organization

## Statements

### Ethics approval and consent to participate

The study was approved by the Ethics Committee of Hospital for Tropical Diseases (approval number 43/HĐĐĐ) and conducted in accordance with the ethical principles of the Declaration of Helsinki.

Consent to participate: not applicable.

### Consent for publication

Not applicable.

### Availability of data and materials

The datasets used and/or analyzed during the current study are available from the corresponding author on reasonable request.

### Authors’ contributions

QMH, BTD, and MCD conceived the study; QMH, BTD, LNTN, TNN, MTN, and KAN collected clinical information; QMH, BTD, NMTB, TTT and MCD analyzed the data; QMH, BTD and MCD interpreted the results; QMH, BTD, TNS, NMTB, TTT, and MCD were major contributors to the writing of the manuscript. All authors contributed to the critical review and editing of the manuscript. All the authors read and agreed with the content.

